# Child mental and behavioral health services during the COVID-19 pandemic: Trends and implications for service outcomes during telehealth expansion

**DOI:** 10.1101/2021.10.10.21264817

**Authors:** Elizabeth N. Riley, Kate D. Cordell, Stephen M. Shimshock, John S. Lyons, Olga A. Vsevolozhskaya

## Abstract

Transportation to/from care is a significant barrier to healthcare access and utilization. The novel coronavirus pandemic prompted a widespread expansion of telehealth service delivery throughout much of 2020. We used propensity score matching to generate two comparison groups of children served in a large public mental and behavioral health system between (1) April-December 2019 (pre-pandemic; n=2,794), and (2) between April-December 2020 (during the COVID-19 pandemic, n=2,794), followed by longitudinal linear mixed-effects modelling to explore the relationship between caregiver transportation needs and child-level outcomes. Our analyses indicated a statistically significant association between the resolution of caregiver’s transportation needs and children’s clinical improvement in the 2019 (pre-pandemic) sample; there was no such association found in the 2020 (pandemic) sample. Our findings suggest that the use of telehealth may mitigate the effect of caregiver transportation needs on child-level clinical outcomes.

## INTRODUCTION

The coronavirus pandemic (COVID-19) has disrupted models of the healthcare delivery system in the United States (US). In person office visits with healthcare providers were limited when stay-at-home orders were imposed, which led to policy changes increasing telehealth coverage by nearly all federal, state, and private insurers.^1^ Healthcare providers began providing services via telehealth, leading to a rapid and massive expansion of this technology during the COVID-19 pandemic.^2^ Telehealth has allowed many patients to continue to access healthcare while mitigating the risk of virus exposure. Multiple reports have investigated racial and ethnic health disparities associated with telehealth use,^3-6^ utilization of telehealth among older adults,^7^ in nursing homes,^8^ and within large healthcare systems.^9,10^ However, there is limited research on the associations between telehealth use and patient-level outcomes. The COVID-19 pandemic greatly increased the urgency and breadth with which telehealth was deployed in the US, and it is possible that the expanded use of telehealth services will remain. There is a need for additional research on telehealth utilization and clinical outcomes, especially among child populations.

Hundreds of thousands of youth and families are served by public mental and behavioral health systems, and many of these children have a complex presentation of medical, psychological, and psychosocial needs. The prevalence rates of US children diagnosed with a mental, behavioral, or developmental disorder are high (approximately 17%),^11^ and mental health concerns in children and adolescents have been rising,^12^ particularly since the onset of the COVID-19 pandemic.^13^ Access to mental and behavioral health services remains a significant challenge, particularly for children living in low-income families and in rural communities.^12,14^ Transportation barriers impact healthcare access, utilization and clinical outcomes for children. Approximately 9% of children living in families with annual incomes <$50,000 miss essential medical appointments due to transportation barriers, regardless of insurance status.^15-17^ Furthermore, children and youth whose caregivers had unmet transportation needs to/from care demonstrate poorer clinical outcomes over time than those youth whose caregivers either never had unmet transportation needs or whose caregivers resolved transportation needs.^18^

Riley et al. demonstrated that a caregiver’s access to transportation is crucial for improvement of child-level health outcomes.^18^ However, the rapid expansion of telehealth during COVID-19 pandemic may lessen the importance of transportation in clinical and functional outcomes. The present study explores how the switch from in-person to telehealth service delivery impacted client-level health outcomes in Idaho’s public mental and behavioral health system using the same data as in Riley et al.^18^ We investigated the relationship between caregiver transportation needs and child-level outcomes in two samples: one sample of children served prior to the start of the pandemic (April-December 2019), and another served during the COVID-19 pandemic when the use of telehealth grew rapidly (April-December 2020).^19^ Consistent with national trends, reported data suggest a huge number of services delivered to children in Idaho’s Department of Behavioral Health were delivered via telehealth during the first year of the pandemic.^20^ We hypothesized the negative impact of caregiver transportation needs on a child’s improvement would be lower in 2020 sample than in the 2019 sample, based on the expansion of telehealth services during the pandemic.

## MATERIALS AND METHODS

### Data sources and study population

Child and adolescent behavioral health data were drawn from the ICANS system, an electronic repository of the Idaho Department of Behavioral Health.^21^ To examine trends in caregiver’s transportation access and youth’s psychosocial improvement, we analyzed behavioral assessments of n = 2,794 children and adolescents (≥5 years of age) that took place between April 1, 2020 and December 31, 2020. March 25, 2020 was the start date of Idaho’s statewide stay-home order,^22^ and COVID-19 vaccines became more widely available starting in January, 2021; this study timeframe corresponded closely with increases in telehealth delivery around the country.^23^ The study was conducted with the approval of the University of Kentucky Institutional Review Board.

### Child and Adolescent Needs and Strengths (CANS) measures

Comprehensive behavioral assessments for children in this sample were performed using the Child and Adolescent Needs and Strengths (CANS).^24,25^ The CANS is a non-diagnostic functional and clinical assessment that has demonstrated adequate reliability when used by both researchers and clinicians^26,27^ and is the most widely used functional assessment for youth served in the public sector in the United States.^28^ The Idaho CANS is administered at the beginning of care and every 90 days during care, and it uses 82 items across six domains: strengths (16 items), life-functioning (14 items), behavioral/emotional needs (18 items), risk behaviors (15 items), and caregiver resources (19 items). The CANS instrument rates each item on a 0 (no need for action) to 3 (immediate action is needed) scale. An item is rated as “actionable” if rated 2 or above, indicating that action is warranted to address the need. Details on the ICANS instrument and methods are available in prior CANS-related publications.^18,29,30^

### Analytic approach

The primary outcomes were defined as in Riley et al.:^18^ the proportion of needs resolved over new needs acquired after the study baseline (during the study timeframe) and the proportion of strengths built over strengths to develop. Preliminary analyses (Supplement Table 1) revealed that while in treatment, there were no appreciable differences in the average number of needs resolved or strengths acquired between the two evaluation periods. However, children in the 2020 sample tended to acquire fewer life-functioning needs (P < 0.001) and risky-behaviors needs (P = 0.002). To ensure our findings were not due to systematic differences in characteristics of children served by the state after the initiation of widespread stay-home orders, we performed a one to one propensity score nearest neighbor matching and obtained data on n = 2,794 children and adolescents (≥5 years of age) assessed between April 1, 2019 and December 31, 2019. Four variables were used as covariates for propensity score matching: (1) the total number of actionable items (TAI) in the strength domain at the initial assessment; (2) TAI in life-functioning domain at initial assessment; (3) TAI in behavioral/emotional needs domain at initial assessment; and (4) TAI in risk behaviors domain at initial assessment.

**Table 1:** Characteristics of study population before and during the COVID-19 pandemic.

|  | April-December,<br>2019<br>n = 2 794 | April-December,<br>2020<br>n = 2 794 | P-value |
| --- | --- | --- | --- |
| Caregiver transportation<br>need |  |  |  |
| No issues | 2 623 (94.2%) | 2 671 (95.6%) | $P = 0.028$ |
| Resolved | 63 (2.3%) | 39 (1.4%) |  |
| Unresolved | 99 (3.5%) | 84 (3.0%) |  |
| LOS (days) | 112 (44.5) | 112 (50.1) | $P = 0.975$ |
| Number of assessments | 2.29 (0.53) | 2.36 (0.59) | $P < 0.001$ |
| Age (years) | 11.7 (3.61) | 12.0 (3.69) | $P = 0.124$ |
| Gender |  |  |  |
| Female | 1 131 (40.5%) | 1 393 (49.9%) | $P = 0.079$ |
| Male | 1 663 (59.5%) | 1 401 (50.1%) |  |
| Race/Ethnicity |  |  |  |
| White | 1 767 (63.2%) | 1 670 (59.8%) | $P < 0.001$ |
| Black/African American | 76 (2.7%) | 93 (3.3%) |  |
| Asian | 313 (11.2%) | 426 (15.2%) |  |
| Hispanic/Latino | 497 (17.8%) | 428 (15.3%) |  |
| Other | 141 (5.1%) | 178 (6.3%) |  |
| Strengths TAI | 3.99 (3.18) | 4.02 (3.16) | $P = 0.733$ |
| Life-functioning TAI | 1.98 (2.01) | 2.01 (2.03) | $P = 0.648$ |
| Behavior-emotional TAI | 3.91 (3.15) | 4.05 (3.20) | $P = 0.110$ |
| Risky-behaviors TAI | 0.84 (1.46) | 0.87 (1.55) | $P = 0.388$ |
| Strengths acquired | 1.27 (2.01) | 1.34 (2.01) | $P = 0.208$ |
| Strengths to develop | 4.86 (3.57) | 5.06 (3.61) | $P = 0.037$ |
| Life-functioning needs<br>resolved | 0.83 (1.45) | 0.88 (1.45) | $P = 0.133$ |
| Life-functioning needs<br>acquired | 2.62 (2.42) | 2.71 (2.49) | $P = 0.182$ |
| Behavior-emotional needs<br>resolved | 1.37 (2.16) | 1.50 (2.26) | $P = 0.117$ |
| Behavior-emotional needs<br>acquired | 4.98 (3.74) | 5.06 (3.78) | $P = 0.436$ |
| Risky-behaviors needs<br>resolved | 0.49 (1.14) | 0.54 (1.25) | $P = 0.131$ |
| Risky-behaviors needs<br>acquired | 1.24 (1.92) | 1.25 (2.04) | $P = 0.834$ |

The main predictor of interest was similarly defined:^18^ caregiver’s transportation needs, defined as “no issues” with transportation needs during the study timeframe, “resolved” transportation needs, and “unresolved” transportation needs. To explore trends in the proportions of needs resolved or strengths acquired during the study timeframe, we fitted a linear mixed-effects model to the aforementioned outcomes and used caregiver’s transportation needs, length of care episode, an interaction between caregiver’s transportation needs and length of care episode, gender, race/ethnicity, and the first five principal components of caregiver’s characteristics as the fixed effects. Under this model, we evaluated an association between caregiver’s transportation needs and children’s proportions of needs resolved (or strengths acquired) over time by assessing statistical significance of the interaction term between child’s length of episode of care and caregiver’s transportation needs. All analyses were performed using R statistical software (version 3.6.3).

## RESULTS

Figure 1 displays dates of the Idaho CANS assessments during the pre-pandemic period and after the statewide stay-home order: between April 1 and December 31, 2020 there was almost a 2-fold reduction in the total number of CANS assessments (n = 6588) relative to assessments conducted during the same timeframe in 2019, prior to the pandemic (n = 10299). Table 1 displays sample characteristics of matched children. Supplementary Table 1 displays pre- and pandemic samples without propensity score matching. Table 1 indicates that in comparison to 2019, fewer caregivers reported transportation need as “resolved” (2.3% vs. 1.4%, *P* = 0.028), children and youth were assessed more frequently (2.29 vs. 2.36 assessments, P<0.001), there were more girls, Asian and White children coming into the system.

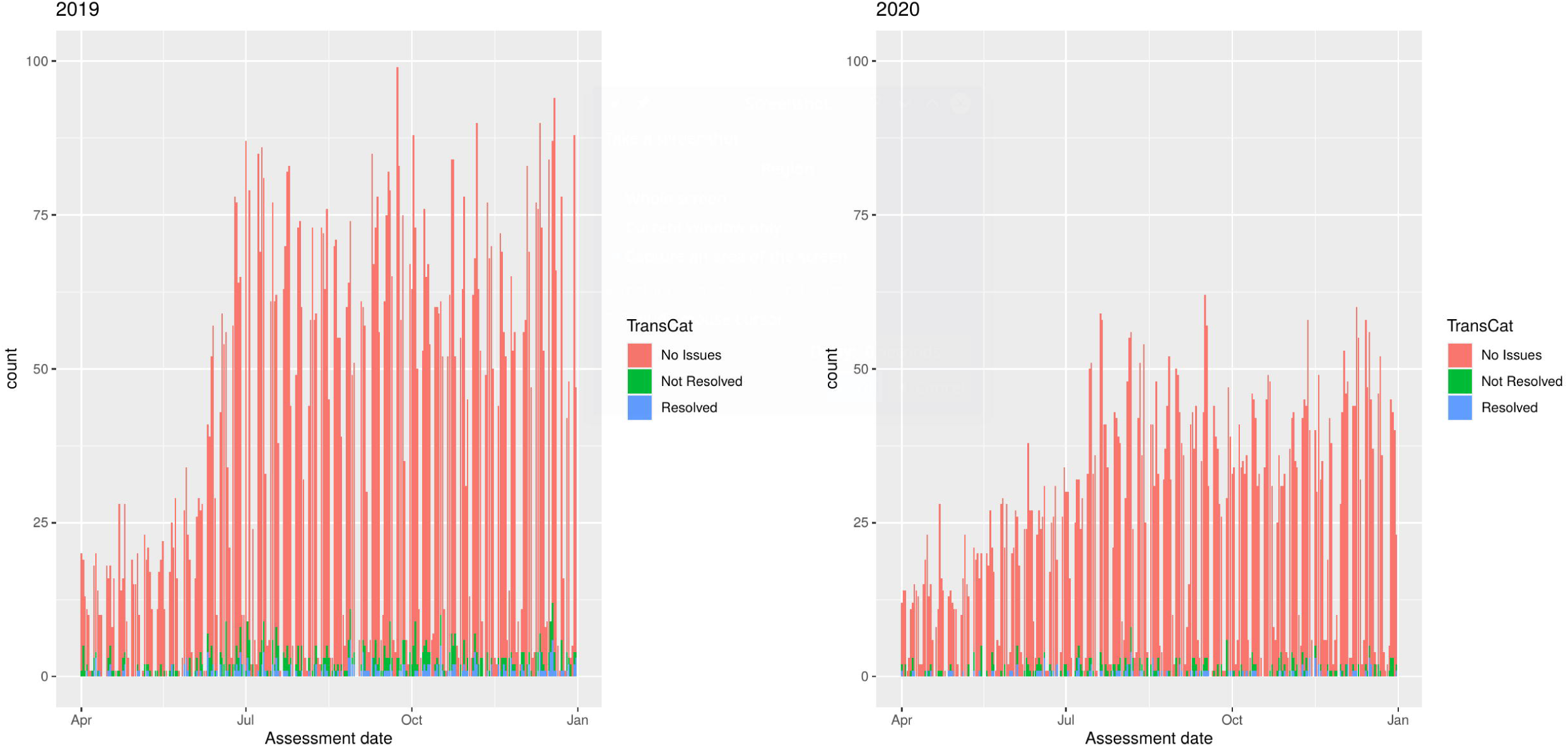

Table 2 shows the results of the linear mixed-model analyses adjusted for gender, race/ethnicity, and caregiver characteristics. The estimated coefficient, 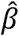, shows percent of areas addressed (i.e., needs resolved or strengths acquired) within each domain associated with every additional 90 days in the system (i.e., between two consecutive CANS assessments) among children whose caregivers never had issues with transportation to/from care. The estimated change, Δ, shows deviations from these percent of addressed areas among children whose caregivers resolved their transportation issues, Δ_*R*_, or did not resolve transportation issues, Δ_*NR*_. Supplementary Table 2 shows the results of the same analyses but on the pre-pandemic sample without propensity score matching.

**Table 2:** Linear mixed-effects model results for the association between areas addressed based on caregiver’s transportation needs.

| CANS Domain | April-December, 2019<br>n = 2,794 |  | April-December, 2020<br>n = 2,794 |  |
| --- | --- | --- | --- | --- |
|  | Resolution rate | P - value | Resolution rate | P - value |
| **Strengths** |  |  |  |  |
| No Issues | $\hat{\beta} = 15.38\%$ | | $\hat{\beta} = 16.01\%$ | |
| Resolved | $\Delta_R = -0.5\%$ | 0.837 | $\Delta_R = 4.2\%$ | 0.227 |
| Not Resolved | $\Delta_{NR} = -5.5\%$ | **0.012\*** | $\Delta_{NR} = -2.5\%$ | 0.313 |
| **Life-functioning** |  |  |  |  |
| No Issues | $\hat{\beta} = 18.56\%$ | | $\hat{\beta} = 18.55\%$ | |
| Resolved | $\Delta_R = 4.88\%$ | 0.138 | $\Delta_R = 6.85\%$ | 0.072 |
| Not Resolved | $\Delta_{NR} = -5.21\%$ | **0.047\*** | $\Delta_{NR} = -5.53\%$ | 0.104 |
| **Behavior-emotional** |  |  |  |  |
| No Issues | $\hat{\beta} = 17.21\%$ | | $\hat{\beta} = 17.89\%$ | |
| Resolved | $\Delta_R = 0.42\%$ | 0.901 | $\Delta_R = 0.76\%$ | 0.842 |
| Not Resolved | $\Delta_{NR} = -6.12\%$ | **0.012\*** | $\Delta_{NR} = -3.82\%$ | 0.148 |
| **Risky-behaviors** |  |  |  |  |
| No Issues | $\hat{\beta} = 23.95\%$ | | $\hat{\beta} = 24.35\%$ | |
| Resolved | $\Delta_R = -0.3\%$ | 0.945 | $\Delta_R = 2.22\%$ | 0.664 |
| Not Resolved | $\Delta_{NR} = -1.82\%$ | 0.618 | $\Delta_{NR} = -4.25\%$ | 0.311 |

Table 2 indicates that, in the pre-pandemic sample, there was a statistically significant association between the resolution of caregiver’s transportation needs and children’s need reduction rate in life-functioning and behavior emotional domains, and in children’s strength acquiring rate. Results without propensity score matching (Supplementary Table 2) are consistent with the findings of Riley et al.^18^ In contrast, we found no statistically significant associations between caregiver’s transportation need resolution and children’s improvement across all four CANS domains in analyses performed using assessments during the COVID-19 pandemic. Additionally, the magnitude of Δ_*NR*_ deviations in strength and behavior-emotional domains in this sample were nearly half those observed in the pre-pandemic sample.

## DISCUSSION

This report demonstrates the impact of COVID-19 on the relationship between the resolution of caregiver transportation needs and children’s strengths/needs development across multiple functioning domains. There was a significant association between resolution of caregiver transportation needs and child-level clinical outcomes pre-pandemic (April-December 2019), such that the resolution of caregiver transportation needs was associated with greater need reduction in the life-functioning domain and greater strength building. Clinically, this represents ∼5% additional needs resolved and 5% additional strengths developed with the resolution of caregiver’s transportation needs. This relationship was not significant, however, when the same analyses were conducted using data during the COVID-19 pandemic (April-December 2020). These results suggest that the expansion of telehealth service delivery may mitigate the impact of caregiver transportation needs on clinical outcomes for children being served in public mental and behavioral health systems.

The results of this study should be contextualized within its limitations. There were no precise service data available that described the exact proportion of services delivered via telehealth vs. those delivered in-person during the pandemic. Data from the Idaho Office of the Governor indicates a 40-fold increase in telehealth sessions from March through May 2020.^20^ However, more data are needed in order to better contextualize the findings and to directly investigate the relationship between caregiver transportation needs, telehealth service delivery, and child-level outcomes.

Substantially fewer children were served in Idaho in 2020 compared to 2019. Matched pre- and during-pandemic samples were largely comparable based on demographic and clinical variables, but the unmatched number of children served and number of CANS assessments done in April-December 2020 was nearly half those during the same time period in 2019. While this reduction in care delivery is consistent with some documented trends of reduced healthcare delivery in the US during the pandemic,^31^ we cannot rule out that some other sample-specific factors mitigated the impact of transportation. Given the remaining large sample size, we can rule out statistical power as a potential threat to the validity of this study.

The intensity and quality of care differences are unknown. It is possible that children served during pandemic received fewer, shorter-length or less organized services. However, results indicated that overall treatment effects (i.e., estimated r’s) did not significantly differ pre- and during pandemic, indicating that the care provided was of measurable value to children served during the pandemic.

Transportation-related needs are one of many barriers to healthcare access, particularly among populations who already have lower access to services.^32^ Clinicians, researchers, and policy makers are advocating for permanent expansion of telehealth services, and both patients and providers report high levels of satisfaction with telehealth.^19^ Telehealth is likely here to stay, but the resolution of caregiver transportation needs should still be an area of clinical intervention for children served in public systems.^18^ Based on these results, one such systems-level change may be supporting and advocating for the continued use of telehealth for clients with transportation needs as a bridge to support service delivery while also working towards the resolution of transportation needs.

## Supporting information

Supplementary results

## Data Availability

All data produced in the present study are available upon reasonable request to the authors

## AUTHOR CONTRIBUTIONS

All authors made substantial contributions to study conception and design, interpretation of study results, writing, reviewing and editing. SMS and OAV performed acquisition of data and statistical analysis.

## SUPPLEMENTARY MATERIAL

I am planning to put result of 2019 data analysis as a supplement.

