## Supplementary results for "Child mental and behavioral health services during the COVID-19 pandemic: Trends and implications for service outcomes during telehealth expansion"

**Supplementary Table 1:** Characteristics of study population before and during the COVID-19 pandemic without propensity score matching.

|  | April-December, 2019  n = 4,458 | April-December, 2020  n = 2,794 | *P*-value |
| --- | --- | --- | --- |
| Caregiver transportation need |  |  |  |
| No issues  Resolved  Unresolved | 4 196 (94.1%) 86 (1.9%) 176 (3.9%) | 2 671 (95.6%) 39 (1.4%) 84 (3.0%) | *P* = 0.024 |
| LOS (days) | 112 (44.5) | 112 (50.1) | *P* = 0.975 |
| Number of assessments | 2.29 (0.53) | 2.36 (0.59) | *P* < 0.001 |
| Age (years) | 11.9 (3.62) | 12.0 (3.69) | *P* = 0.124 |
| Gender  Female  Male | 2 127 (47.7%) 2 331 (52.3%) | 1 393 (49.9%) 1 401 (50.1%) | *P* = 0.079 |
| Race/Ethnicity  White  Black/African American  Asian  Hispanic/Latino  Other | 2 806 (62.9%)  142 (3.2%)  508 (11.4%)  767 (17.2%)  235 (5.3%) | 1 670 (59.8%)  93 (3.3%)  426 (15.2%)  428 (15.3%)  178 (6.3%) | *P* = 0.157 |
| Strengths acquired | 1.37 (2.13) | 1.34 (2.01) | *P* = 0.467 |
| Strengths to develop | 5.28 (3.59) | 5.06 (3.61) | *P* = 0.011 |
| Life-functioning needs resolved | 0.91 (1.45) | 0.88 (1.45) | *P* = 0.433 |
| Life-functioning needs acquired | 2.93 (2.51) | 2.71 (2.49) | *P* < 0.001 |
| Behavior-emotional needs resolved | 1.42 (2.21) | 1.50 (2.26) | *P* = 0.117 |
| Behavior-emotional needs acquired | 1.40 (2.02) | 1.25 (2.04) | *P* = 0.142 |
| Risky-behaviors needs resolved | 0.55 (1.19) | 0.54 (1.25) | *P* = 0.664 |
| Risky-behaviors needs acquired | 1.40 (2.02) | 1.25 (2.04) | *P* = 0.002 |

**Supplementary Table 2**: Linear mixed-effects model results for the association between needs-resolving rate over time based on caregiver’s transportation needs without propensity score matching.

| CANS Domain | April-December, 2019  n = 4,458 | | April-December, 2020  n = 2,794 | |
| --- | --- | --- | --- | --- |
|  | Resolution rate | *P* - value | Resolution rate | *P* - value |
| **Strengths**  No Issues  Resolved  Not Resolved | $\hat{\beta}=15.27\%$  $\Delta_{R}=1.61\%$  $\Delta_{NR}=-4.26\%$ | $0.482$  ${\mathbf{0.0}\mathbf{09}}^{\boldsymbol{*}}$ | $\hat{\beta}=16.01\%$  $\Delta_{R}=4.2\%$  $\Delta_{NR}=-2.5\%$ | $0.227$  $0.313$ |
| **Life-functioning**  No Issues  Resolved  Not Resolved | $\hat{\beta}=18.60\%$  $\Delta_{R}=4.58\%$  $\Delta_{NR}=-4.56\%$ | $0.094$  ${\boldsymbol{0.0}\boldsymbol{21}}^{*}$ | $\hat{\beta}=19.56\%$  $\Delta_{R}=7.48\%$  $\Delta_{NR}=-5.14\%$ | $0.072$  $0.084$ |
| **Behavior-emotional**  No Issues  Resolved  Not Resolved | $\hat{\beta}=16.85\%$  $\Delta_{R}=2.44\%$  $\Delta_{NR}=-4.39\%$ | $0.638$  $\boldsymbol{0.016}\boldsymbol{*}$ | $\hat{\beta}=18.47\%$  $\Delta_{R}=6.2$%  $\Delta_{NR}=-3.94\%$ | $0.878$  $0.152$ |
| **Risky-behaviors**  No Issues  Resolved  Not Resolved | $\hat{\beta}=24.20\%$  $\Delta_{R}=0.9\%$  $\Delta_{NR}=-1.65\%$ | $0.796$  $0.542$ | $\hat{\beta}=26.84\%$  $\Delta_{R}=8.09$%  $\Delta_{NR}=-6.32\%$ | $0.174$  $0.176$ |
